# Shannon Entropy Trajectories Reveal Between-Arm Distributional Structure Invisible to Standard Endpoint Analysis in Pooled ALS Clinical Trials

**DOI:** 10.64898/2026.04.20.26351319

**Authors:** Anderson M. Rodriguez, the Pooled Resource Open-Access ALS Clinical Trials Consortium

**Author notes:** Data used in the preparation of this article were obtained from the Pooled Resource Open-Access ALS Clinical Trials (PRO-ACT) Database. As such, the following organizations and individuals within the PRO-ACT Consortium contributed to the design and implementation of the PRO-ACT Database and/or provided data, but did not participate in the analysis of the data or the writing of this report: Alexion Pharmaceuticals, Inc.; ALS Therapy Alliance; Amylyx Pharmaceuticals, Inc.; Apellis Pharmaceuticals, Inc.; Cytokinetics, Inc.; Knopp Bio-sciences; Neuraltus Pharmaceuticals, Inc.; Neurological Clinical Research Institute, MGH; Northeast ALS Consortium; Novartis; Orion Corporation; Prize4Life Israel; Regeneron Pharmaceuticals, Inc.; Sanofi; Teva Pharmaceutical Industries, Ltd.; The ALS Association; The Sean M. Healey & AMG Center for ALS at Massachusetts General Hospital.

## Abstract

Standard analysis of amyotrophic lateral sclerosis (ALS) clinical trials evaluates therapeutic efficacy by comparing linear slopes of total ALS Functional Rating Scale (ALSFRS) scores between treatment arms. This approach compresses multidomain ordinal data into a single scalar trajectory, discarding distributional structure. When subgroup-level trends differ in timing or direction, such aggregation can attenuate or eliminate them, a phenomenon known as Simpson’s paradox. Here we apply Shannon entropy, computed from item-level score distributions within each ALSFRS functional domain following the framework established in [8], to the PRO-ACT database, stratified by treatment arm (Active: *n* = 4,581; Placebo: *n* = 2,931; 19 monthly time points). The entropy trajectories of drug-treated and placebo populations diverge visibly and systematically across all four functional domains (Bulbar, Fine Motor, Gross Motor, Respiratory). In the Fine Motor domain, the placebo population reaches peak entropy at month 8 and reverses, while the active population does not peak until month 13, a five-month delay in the population’s transit toward functional loss. This divergence is model-independent: it is present in the raw Shannon entropy trajectories before any dynamical model is applied. A permutation test shuffling patient-level arm labels (*n* = 1,000 permutations) confirms that the total integrated absolute divergence across all four domains exceeds the null distribution at *p <* 0.001 (observed: 4.48; null: 2.03 *±* 0.33; 7.5 standard deviations above the null mean), with Fine Motor (*p* = 0.001) and Respiratory (*p <* 0.001) individually significant. The quantity that differs between arms, the shape and timing of the population’s distributional evolution, does not exist as a measurable quantity in the total-score linear-slope framework used to evaluate these trials. Whether this signal reflects genuine treatment effects, compositional artifacts from pooling heterogeneous trials, or both cannot be determined from the anonymized public database alone. What can be determined is that the standard ALS clinical trial endpoint makes an implicit assumption, that the distributional information it discards is uninformative, and the present results demonstrate empirically that this assumption is false.

## 1 Introduction

The standard analytic pipeline for ALS clinical trials evaluates therapeutic efficacy by summing item-level scores on the ALS Functional Rating Scale (ALSFRS or ALSFRS-R) into a total score, fitting a linear model to the resulting trajectory, and comparing slopes between treatment and placebo arms [1, 2]. This field-standard pipeline involves two stages of information compression: item summation collapses the multidomain structure of the scale into a single scalar, and linear slope estimation discards any nonlinear dynamics in the trajectory.

The consequences of this compression are well documented: the ALSFRS assesses four functionally distinct domains (Bulbar, Fine Motor, Gross Motor, and Respiratory), each of which progresses at different rates and through different mechanisms [3]. Rasch analysis has confirmed the multidimensionality of the ALSFRS-R, concluding that, rather than a single total score, the scale should be treated as a domain profile[4, 5]. A patient whose bulbar function deteriorates while motor function is preserved, for example, can produce the same total-score slope as a patient declining uniformly. In practice, this can lead to the different functional capacities on different outcome trajectories being treated as identical by the standard endpoint analysis approaches [5], i.e., some non-trivial amount of relevant collected information is discarded before analysis ever takes place.

This is a well-known statistical phenomenon, as when subgroup-level trends differ in timing or direction, aggregating across subgroups can attenuate or eliminate the individual trends. The effect is called Simpson’s paradox (discovered first by Pearson (1899), then Yule (1903)[6], before being canonically demonstrated by Simpson in 1951[7]). The phenomenon is a property of the aggregation operator as opposed to a bias in the data. The present study demonstrates that the total-score endpoint performs exactly this lossy aggregation: it sums across four domains that peak, reverse, and compress on different timelines. If a treatment effect operates on domain-specific timing rather than on the rate of uniform decline, the summation step will cancel it.

The present work asks whether this cancellation has occurred in the PROACT dataset[2]. Specifically: does the distributional information *discarded* by the standard endpoint contain non-trivial structure that differs between treatment arms? To address this, we apply the Shannon entropy preprocessing framework established in [8] to the ALSFRS data in the PRO-ACT database [2], stratified by treatment arm. Rodriguez (2026) demonstrated that Shannon entropy, computed from the population-level frequency distribution of item scores within each functional domain at each monthly time point of a Swedish twin aging study cohort (SATSA)[9], provides a model-independent measure of distributional structure. Unlike the total score, entropy preserves information about *how* scores are distributed across categories.

The central finding is that these trajectories diverge systematically across all four functional domains, with the clearest example in Fine Motor, where the drug-treated population’s distributional transition is delayed relative to placebo by several months. This cross-domain divergent signal is, by construction, invisible to total-score linear-slope analysis. This work demonstrates that the quantity which differs between arms (the shape and timing of the population’s distributional evolution) does not exist in the output space of that model. A linear slope of summed scores cannot represent a five-month shift in peak distributional spread. The aggregation step eliminates these features before the slope is ever computed.

The implications are twofold. First, the standard endpoint framework makes an implicit assumption: that the distributional information it discards during item summation is uninformative. The present results demonstrate empirically that this assumption is false. Second, because the PRO-ACT database pools anonymized trials with heterogeneous compounds and enrollment criteria, the source of the signal (whether treatment effects, compositional confounds arising from the pooling of heterogeneous trials, or both) cannot be resolved from the available data. The same Simpson’s paradox mechanism that could mask a real treatment effect during item summation could also create a spurious between-arm difference during trial pooling. What can be determined is that the information the standard endpoint discards is not empty. Reexamination of the individual trials comprising the PRO-ACT dataset is warranted.

## 2 Materials and Methods

### 2.1 Data source

Data were obtained from the Pooled Resource Open-Access ALS Clinical Trials (PRO-ACT) database [2]. The data available in the PRO-ACT Database have been volunteered by PRO-ACT Consortium members. The version used here (accessed 25 March 2026) contains 81,229 records from 9,149 unique patients. After standard cleaning (removing records with missing time stamps or no valid item-level scores, restricting to 18 months from baseline), the analysis dataset comprises 77,889 observations from 8,819 unique patients.

Treatment arm assignments were obtained from the PRO-ACT Treatment Group file and merged on patient identifier. Of the 8,819 patients in the cleaned ALSFRS dataset, 7,512 (85.2%) had treatment arm labels: 4,581 assigned to Active arms and 2,931 to Placebo arms across the constituent trials. The PROACT database anonymizes trial and compound names; the “Active” label pools all experimental compounds across all contributing trials.

### 2.2 ALSFRS domain structure

The 10 ALSFRS items are grouped into four functional domains:

- **Bulbar**: Q1 Speech, Q2 Salivation, Q3 Swallowing
- **Fine Motor**: Q4 Handwriting, Q5 Cutting Food, Q6 Dressing and Hygiene
- **Gross Motor**: Q7 Turning in Bed, Q8 Walking, Q9 Climbing Stairs
- **Respiratory**: Q10 Respiratory function (single item)

Each item is scored on an ordinal scale from 0 (total loss of function) to 4 (normal function), yielding a 5-category discrete distribution[1]; the data share the ordinal structure to which the ECTO preprocessing framework was originally applied[8].

The PRO-ACT database records Fine Motor item Q5 in two variants: Q5a (Cutting Food, without gastrostomy) and Q5b (Cutting Food, with gastrostomy)[2]. Because these variants use different scoring criteria and Q5a covers the large majority of observations (74,348 of 81,229 rows, 91.5%), only Q5a is used in the present analysis. Patients with no Q5a data at a given time point do not contribute to Fine Motor domain entropy at that time point but remain in the analysis for the other three domains. Approximately 0.17% of item-level scores in the PRO-ACT data contain fractional values (e.g., 2.5), likely reflecting trial-specific scoring conventions; these are rounded to the nearest integer before frequency tabulation (half-values rounded to even).

### 2.3 Shannon entropy computation

Following the information-theoretic preprocessing protocol established in Rodriguez (2026)[8], Shannon entropy [10] is computed for each item at each monthly time point from the population-level frequency distribution of scores:

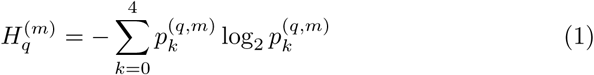

where 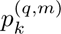 is the relative frequency of score *k* for item *q* at month *m*, and terms with *p_k_* = 0 are omitted by convention. Domain-level entropy is computed as the arithmetic mean of item-level entropies within each domain:

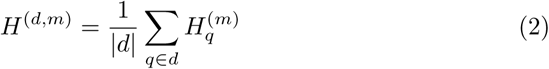

Because the PRO-ACT data are monthly and subject to attrition and heterogeneous visit schedules, individual item-month bins can become sparse. A minimum of 30 observations per item per month is therefore required for inclusion, ensuring at least six observations per score category on average for stable frequency estimation. This threshold was not required in the original ECTO application, where each survey wave contained hundreds of respondents[8]; it is introduced here to guard against degenerate frequency distributions at low-count time points. Results are robust to minimum-*n* thresholds of 20 and 50 (Supplementary Table S2).

Visit times are converted to months by dividing days from baseline by 30.44 and rounding to the nearest integer. This procedure yields 19 common time points (months 0–18) for both treatment arms across all four domains.

#### Critically, the entropy computation is performed separately for each treatment arm

At each month, the score distribution for Active patients and the score distribution for Placebo patients are computed independently, and entropy is calculated from each. The resulting per-arm entropy trajectories are the primary data objects of this study.

### 2.4 Divergence metrics

Three model-independent metrics quantify the separation between arm-level entropy trajectories:

1. **Integrated Absolute Divergence (IAD)**: For each domain, the area between the Active and Placebo entropy curves, computed via trapezoidal integration[11] of 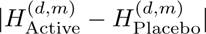 over months 0–18.
2. **Peak timing difference**: The month at which each arm’s entropy trajectory reaches its maximum value, computed independently per domain. A difference in peak month indicates that the two populations reach maximum distributional spread at different times.
3. **Total IAD**: The sum of per-domain IAD values, providing an omnibus measure of trajectory separation.

### 2.5 Permutation test

Statistical significance is assessed by a permutation test. Patient-level arm labels are shuffled (preserving within-patient consistency across time points), entropy trajectories are recomputed for each shuffled arm, and the divergence metrics are recalculated. This is repeated 1,000 times to construct a null distribution. The *p*-value is the proportion of permutations producing divergence at least as large as the observed value. This test is model-independent; as such, it involves no curve fitting, no ODE integration, and no parameter estimation.

## 3 Results

### 3.1 Entropy trajectories diverge between treatment arms

Figure 1 shows the Shannon entropy trajectories for all four ALSFRS functional domains, stratified by treatment arm. The two populations diverge visibly in every domain.

**Figure 1:**
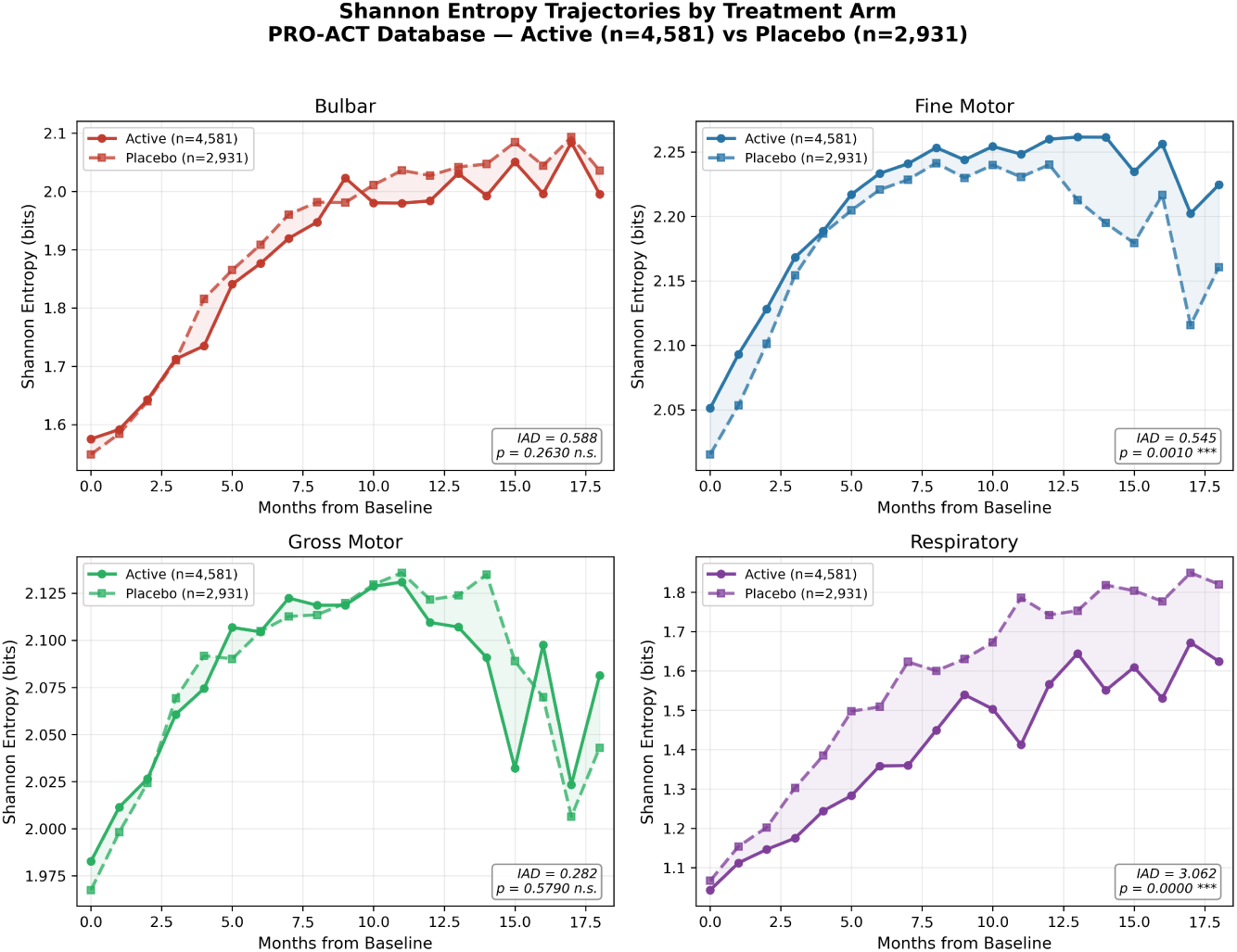
Shannon entropy trajectories for the four ALSFRS functional domains, stratified by treatment arm (Active: *n* = 4,581, solid lines; Placebo: *n* = 2,931, dashed lines). Domain-level entropy is the arithmetic mean of item-level entropies; months 0–18 from baseline. Shaded regions indicate the area between curves. Inset annotations report the integrated absolute divergence (IAD) and permutation test *p*-value for each domain. Fine Motor (*p* = 0.001) and Respiratory (*p <* 0.001) show statistically significant divergence.

In the **Bulbar** domain, both arms rise from approximately 1.55 bits at baseline. The Placebo arm reaches higher entropy values from month 5 onward, peaking near 2.10 bits, while the Active arm plateaus around 2.00 bits by month 10.

In the **Fine Motor** domain, both arms rise from approximately 2.05 bits at baseline (Figure 2). The Placebo arm peaks at month 8 (approximately 2.25 bits) and reverses, declining to approximately 2.16 bits by month 18. The Active arm peaks at month 13 (approximately 2.25 bits) and shows minimal reversal by month 18 (0.037 bits, compared to 0.081 bits in Placebo). The five-month delay in peak timing indicates that the drug-treated population reaches maximum distributional spread, the point at which patients are maximally dispersed across all functional levels, five months later than placebo.

**Figure 2:**
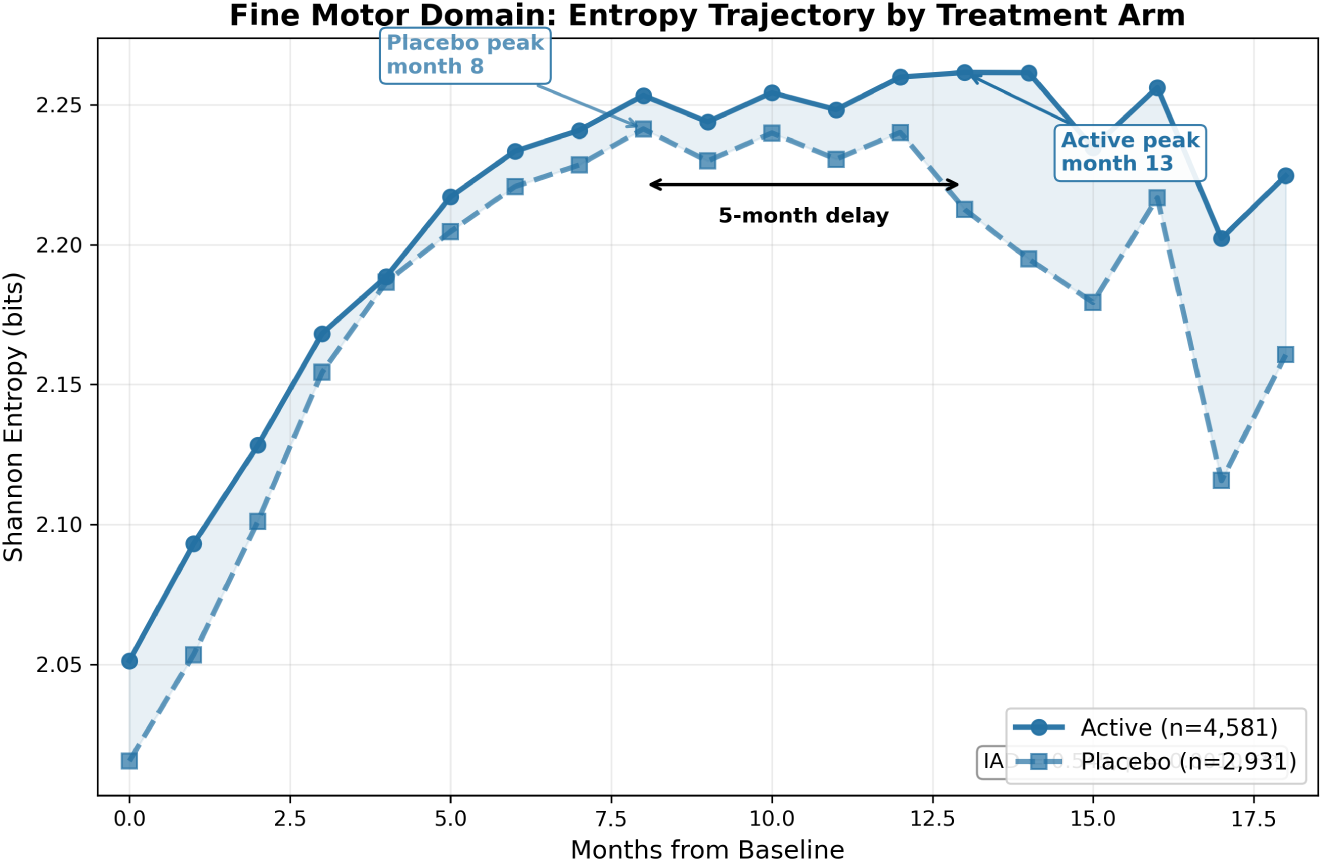
Fine Motor domain entropy trajectories by treatment arm. The Placebo arm peaks at month 8 and reverses; the Active arm does not peak until month 13, a five-month delay in the population’s transit through maximal distributional spread. Shaded region indicates divergence between arms. IAD = 0.545, *p* = 0.001.

In the **Gross Motor** domain, both arms show rise-peak-reversal dynamics. The trajectories are closer together but the Active arm maintains slightly lower entropy at late time points.

In the **Respiratory** domain, the Placebo arm rises faster and reaches higher entropy values from month 5 onward, with the Active arm consistently lower (Figure 3).

**Figure 3:**
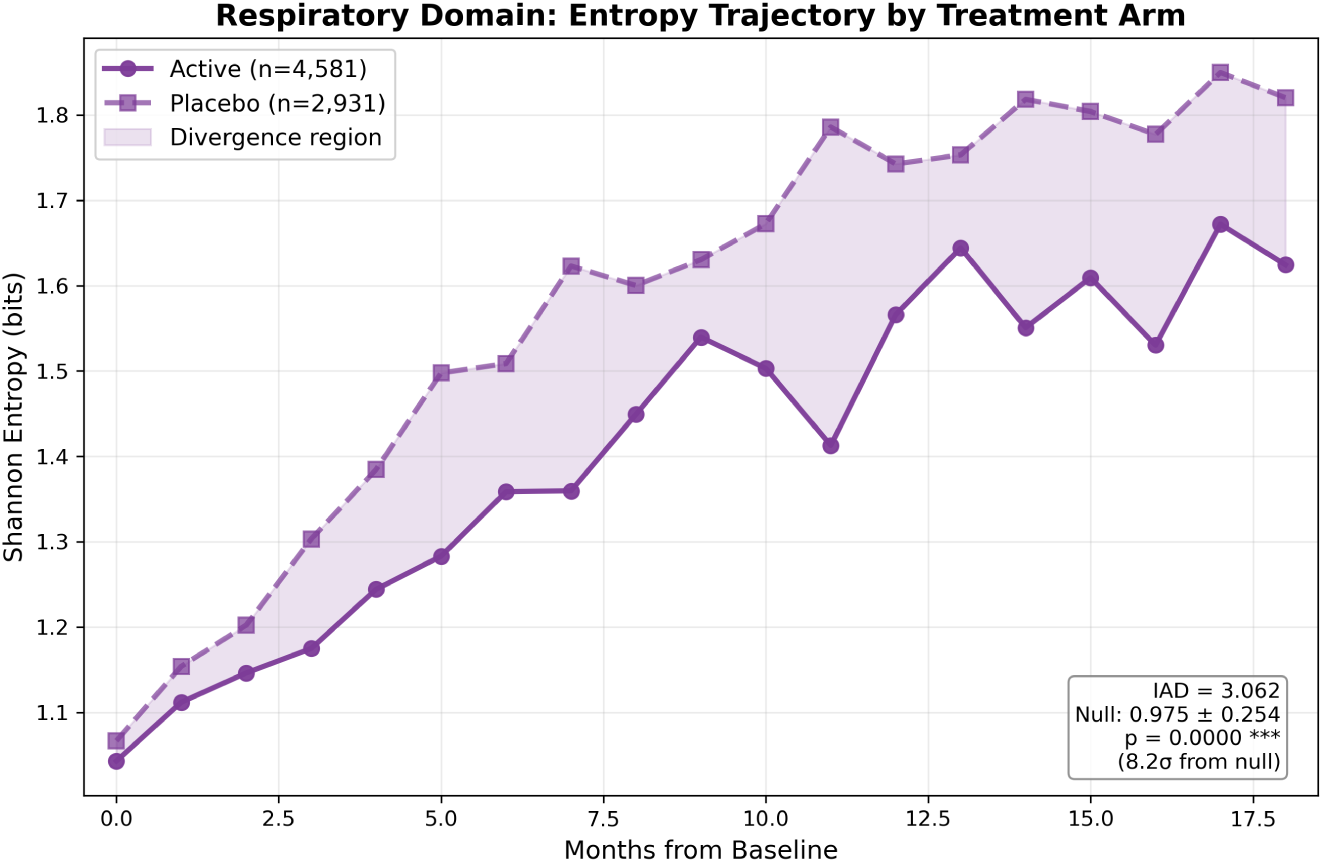
Respiratory domain entropy trajectories by treatment arm. The Placebo arm rises faster and reaches consistently higher entropy from month 4 onward, indicating earlier and greater distributional spread across respiratory function levels. Shaded region indicates the divergence area. IAD = 3.062, *p <* 0.001 (8.2 standard deviations above the permutation null mean).

The direction of the divergence is consistent across all four domains: the Placebo arm reaches higher entropy (greater distributional spread) earlier, while the Active arm evolves through the distributional transition more slowly.

### 3.2 Permutation test

Table 1 and Figure 4 present the results of the permutation test (1,000 permutations, patient-level arm label shuffling). The total integrated absolute divergence across all four domains is 4.48, compared to a null distribution of 2.03 *±* 0.33 (*p <* 0.001). Not one of the 1,000 random arm shuffles produced total divergence as large as the observed value; the observed total IAD lies 7.5 standard deviations above the null mean.

**Figure 4:**
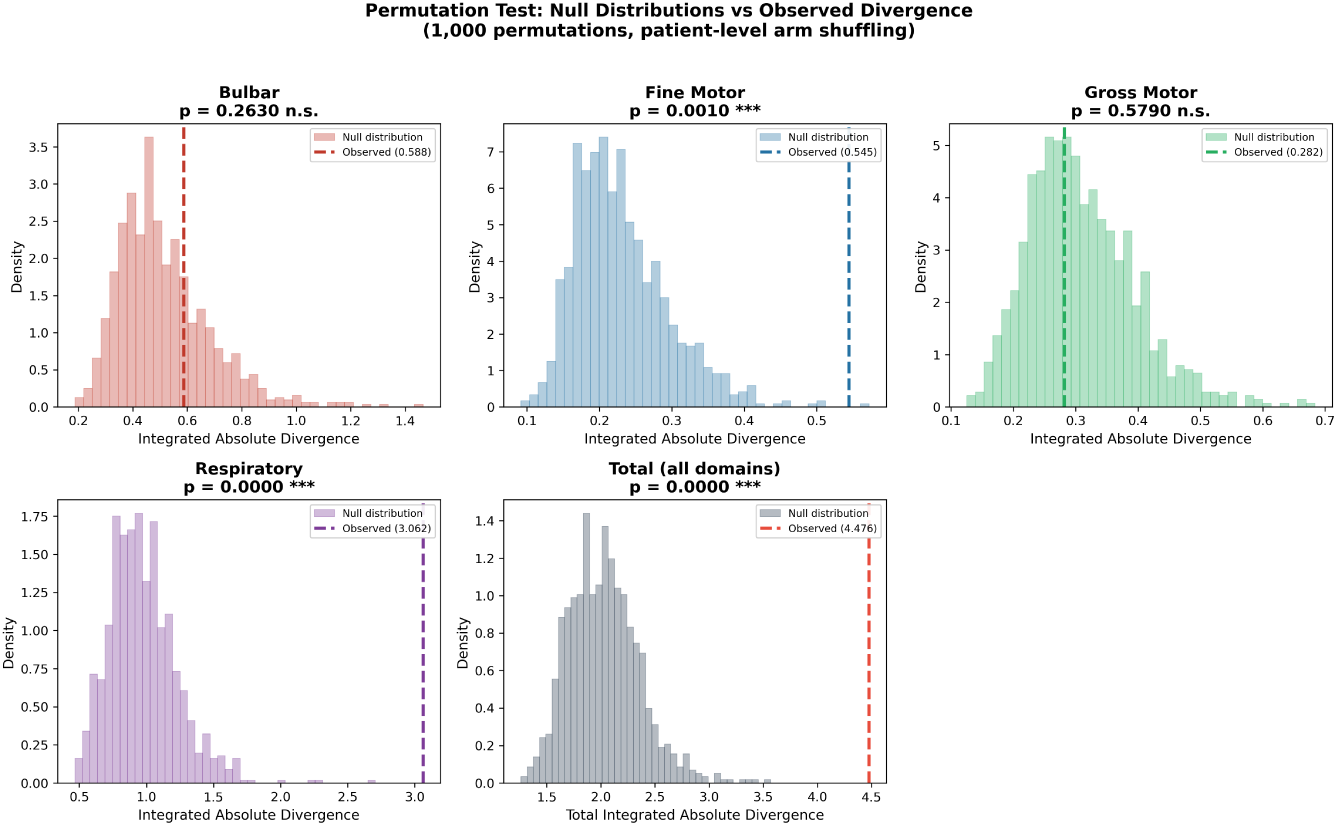
Permutation test null distributions (1,000 permutations, patient-level arm shuffling) and observed integrated absolute divergence (dashed lines) for each ALSFRS functional domain and for the total across all domains. Fine Motor (*p* = 0.001) and Respiratory (*p <* 0.001) show observed values far exceeding the null. The total IAD of 4.48 lies 7.5 standard deviations above its null mean of 2.03 *±* 0.33.

**Table 1:** Permutation test results: integrated absolute divergence (IAD) between Active and Placebo entropy trajectories. 1,000 permutations with patient-level arm label shuffling.

| Domain | Observed IAD | Null $\mu$ | Null $\sigma$ | $p$-value |
| --- | --- | --- | --- | --- |
| Bulbar | 0.588 | 0.514 | 0.169 | 0.263 |
| Fine Motor | 0.545 | 0.230 | 0.065 | 0.001*** |
| Gross Motor | 0.282 | 0.309 | 0.086 | 0.579 |
| Respiratory | 3.062 | 0.975 | 0.254 | <0.001*** |
| Total (sum) | 4.476 | 2.029 | 0.327 | <0.001*** |

At the individual domain level, Fine Motor (*p* = 0.001) and Respiratory (*p <* 0.001) are independently significant (Figure 5). Respiratory shows the largest single-domain effect: an observed IAD of 3.06 against a null of 0.98 *±* 0.25, representing 8.2 standard deviations from the null mean. The numerical dominance of the Respiratory IAD reflects in part the single-item structure of that domain (Q10 alone), where distributional shifts translate directly into entropy changes without averaging across multiple items. Bulbar (*p* = 0.263) and Gross Motor (*p* = 0.579) are individually non-significant but contribute to the total measure.

**Figure 5:**
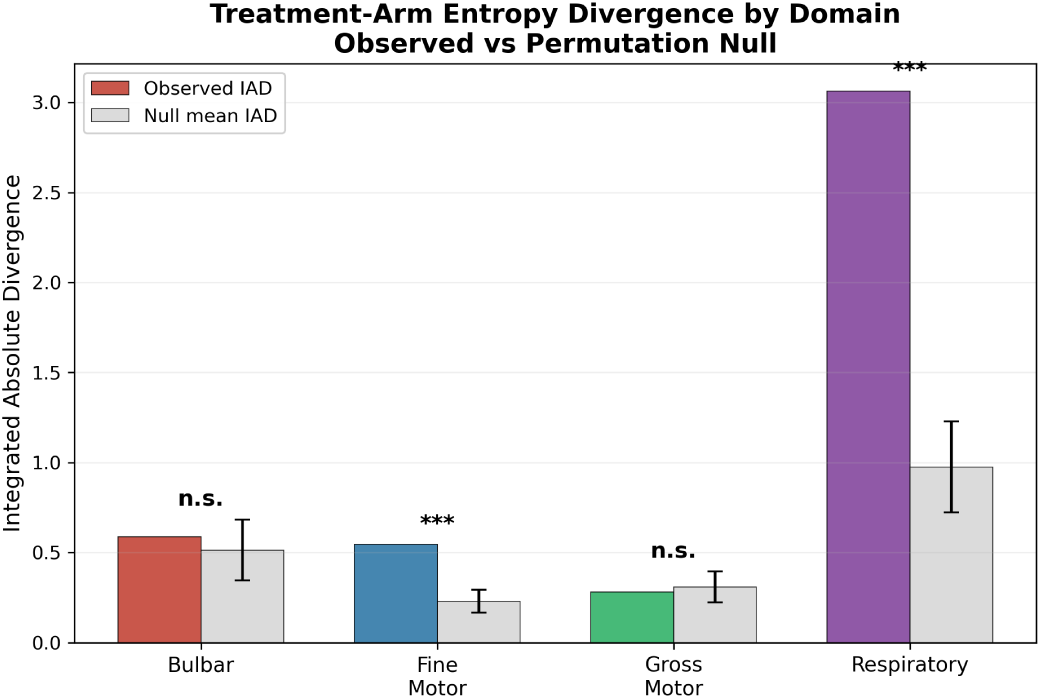
Integrated absolute divergence (IAD) between Active and Placebo entropy trajectories for each ALSFRS functional domain. Colored bars: observed IAD; gray bars: permutation null mean (*±*1*σ*). Fine Motor and Respiratory are individually significant (*p ≤* 0.001); Bulbar and Gross Motor are not.

The Fine Motor time-averaged signed separation is also significant (Δ = 0.031 bits, *p <* 0.001), confirming that the active arm’s entropy trajectory is systematically higher (broader distributional spread maintained longer) than placebo in this domain. The Respiratory time-averaged signed separation is *−*0.167 bits (*p <* 0.001), indicating that the placebo arm reaches higher entropy (greater distributional spread) than the active arm in the respiratory domain.

The five-month peak timing difference in Fine Motor yields *p* = 0.083 under the same permutation procedure, which is suggestive but not individually significant at conventional thresholds. The integrated trajectory separation, which captures the full shape of the divergence rather than a single landmark, is the more powerful measure.

## 4 Discussion

### 4.1 Model-independent frame

The central result of this study, that drug-treated and placebo populations follow visibly different entropy trajectories across all four ALSFRS functional domains, requires no dynamical model, no curve fitting, and no parameter estimation. It follows directly from computing Shannon entropy on score distributions stratified by treatment arm.

This parsimony in calculation is important as it means the finding cannot be attributed to model flexibility, overfitting, or optimizer sensitivity. Since Shannon entropy is a deterministic function of the score distribution, identical results can be achieved given the same data, the same stratification, and the same entropy formula.

### 4.2 On the meaning of divergence

In this study Shannon entropy measures the distributional spread of scores across categories at each time point. A population with high entropy has patients distributed across all functional levels; a population with low entropy has patients clustered together, either at the high-function or low-function end of the scale.

ALS progresses relentlessly from onset[12]: functional scores decline over time, though the rate and pattern of decline vary across patients and domains[13]. At any given time, the population is a mixture of patients at different stages of this decline. Entropy is highest when patients are maximally dispersed across all functional levels: some still near-normal, some in the middle, some near total loss. Entropy falls when the distribution begins to compress, as enough patients have reached the low end of the scale that the population piles up there.

The entropy peak is a tipping point. Before the peak, more patients are entering the middle of the distribution than leaving it; the wave of decline is spreading. After the peak, more patients are leaving the middle toward total loss than entering from the top; the wave is compressing. In the Fine Motor domain, the placebo population hits this tipping point at month 8. The drug-treated population does not hit it until month 13. The active arm delays this tipping point by five months.

This divergence is consistent with a treatment effect, though compositional confounds from pooling heterogeneous trials cannot be excluded (see Limitations). Regardless of its source, this signal is visible in all four domains, consistent in direction, and present across the majority of the observation window.

### 4.3 Standard endpoint analysis cannot detect this signal

The total-score linear-slope analysis asks a single question: does the summed score decline more slowly in the drug arm? This is one dimension of a multidimensional phenomenon. The entropy analysis reveals that the between-arm divergence operates on distributional shape and timing, and not simply on the rate of score decline.

This demonstrates Simpson’s paradox in action. Each domain has its own trajectory with its own peak, reversal, and compression timeline. Summing across domains cancels the timing differences: a five-month delay in the Fine Motor peak is averaged against Respiratory and Gross Motor and Bulbar, each on their own schedule. The resulting total-score slope reflects a weighted average of partially canceling trends, i.e., a drug that shifts domain-specific timing without changing the aggregate rate of decline will produce a negligible difference in the total-score slope and a large, visually obvious difference in the domain-level entropy trajectories.

The limitation is not one of statistical power, but of structure: the signal documented in this study does not exist as a measurable quantity in the output space of the standard model.

### 4.4 Implications for the standard endpoint framework

The PRO-ACT database is composed of completed phase II/III trials, the majority of which did not lead to drug approval [2]. The pooled Active arm in this database represents patients who received experimental compounds that were judged to have no therapeutic effect. The present analysis reveals a systematic, four-domain entropy trajectory divergence between these patients and the placebo controls.

This finding does not permit the conclusion that any specific drug worked. The PRO-ACT database anonymized compounds, and as discussed in the Limitations, the pooling of heterogeneous trials introduces a Simpson’s paradox risk: between-trial differences in phenotype composition and randomization ratios could produce arm-level distributional divergence unrelated to treatment. Resolving this requires individual-trial analysis with unblinded records, which are not, to the author’s knowledge, publicly available.

What the finding does establish is that the standard endpoint framework operates on an empirically false assumption. The total-score linear-slope model implicitly assumes that the distributional information discarded during item summation (how scores are spread across categories within each functional domain) is uninformative for detecting treatment-arm differences. Whether that structure reflects treatment effects, compositional artifacts, or both, *the assumption that it is empty is refuted*.

This concern is not specific to ALS. Any clinical trial that evaluates therapeutic efficacy by summing ordinal items and fitting a linear slope is subject to the same structural limitation. The UPDRS (Parkinson’s disease), ADAS-Cog (Alzheimer’s disease), EDSS (multiple sclerosis), HAM-D (depression), and PANSS (schizophrenia) all share the same structure of ordinal items grouped into domains [14, 15, 16, 17, 18]. The same question applies to all of them: does the distributional information discarded by item summation contain non-trivial structure, and if so, what does it mean?

### 4.5 Limitations

Several limitations should be noted.

#### Compound heterogeneity

The PRO-ACT database de-identifies trial and compound names; the pooled Active arm may contain drugs with heterogeneous mechanisms, and the observed effect could be driven by a subset of compounds. Decomposing the signal to individual drugs requires the unblinded trial records, which are not publicly available.

#### Phenotype heterogeneity

ALS encompasses distinct progression phenotypes (bulbar-onset versus limb-onset, fast versus slow progressors) that may be distributed unevenly across the constituent trials and, consequently, across the pooled Active and Placebo arms. If one trial enrolled predominantly fast progressors and used a 2:1 Active:Placebo ratio while another enrolled slow progressors at 1:1, the pooled arms would differ in phenotype composition for reasons unrelated to treatment. The permutation test shuffles patient-level arm labels across the entire pooled dataset, destroying any trial-level structure, and therefore cannot separate treatment effects from systematic between-trial differences in phenotype composition. This is an aggregation-induced compositional confound inherent to any pooled analysis of heterogeneous trials.

### Population-level analysis

The entropy analysis operates at the population level and does not provide individual patient predictions. It detects shifts in the population’s distributional evolution, not individual trajectories.

#### Differential dropout

Survivor bias may contribute to late-timepoint divergence, though the consistency across domains and the early onset of divergence (by month 4–5) mitigate this concern. Supplementary Table S1 and Figure S1 report the number of patients contributing observations at each monthly time point by treatment arm. The permutation test addresses this concern to the extent that arm labels are shuffled at the patient level while preserving each patient’s complete observation pattern, including their dropout time. The null distribution therefore captures the range of entropy trajectory divergence achievable under random arm assignment, including any divergence arising from chance compositional asymmetries in dropout. The significance of the observed divergence (*p <* 0.001) establishes that the signal exceeds what random label assignment produces. However, the permutation test cannot fully disentangle a direct treatment effect on score distributions from an indirect compositional effect mediated by treatment-dependent dropout: if the drug prolongs survival, the composition of who is observed at late time points differs systematically between arms in a way that random shuffling does not reproduce. This remaining confound does not undermine the finding (differential survival *is itself a treatment effect*) but leaves open whether the entropy divergence reflects altered disease dynamics, altered observation composition, or both.

#### Visit scheduling heterogeneity

The PRO-ACT database pools trials with different visit schedules (some monthly, some bimonthly, some quarterly). When visit times are rounded to the nearest integer month, the number of patients contributing observations at a given month varies non-monotonically (Supplementary Figure S1), reflecting the mixture of scheduling protocols rather than true dropout and re-entry. This creates months with higher and lower sample sizes. Because entropy is estimated from empirical frequency distributions, months with fewer observations produce noisier estimates. The minimum-*n* threshold of 30 per item per month provides a floor, but entropy precision still varies with sample size. Critically, this visit-density variation affects both arms equally (the same pooled trials contribute patients to both Active and Placebo) so the between-arm *difference* is less affected than the individual trajectories. The permutation test, which preserves each patient’s observation pattern, inherits the same visit-density structure in both observed and null conditions.

#### Instrument scope

The analysis uses the original ALSFRS scale (Q1–Q10), not the revised ALSFRS-R. The Respiratory domain is represented by a single item (Q10); patients assessed with the ALSFRS-R respiratory subscale (R1–R3) contribute to the other three domains but not to Respiratory. Fine Motor uses Q5a (without gastrostomy) only; coalescing Q5a and Q5b would mix distinct scoring instruments into a single frequency distribution, which is inconsistent with the information-preserving approach motivating this study.

#### Confounders

The permutation test does not adjust for potential confounders such as site effects or baseline severity imbalances, which are partially addressed by the randomization within constituent trials.

### 4.6 Future directions

A logical extension of this framework is individual-trial decomposition. The observed signal is pooled across de-identified trials with unknown compounds; it cannot be attributed to any specific drug from the public database alone. Decomposing the entropy trajectories by individual trial would determine whether the between-arm divergence is concentrated in a subset of compounds or distributed across the full set, and would resolve the Simpson’s paradox concern identified in the Limitations. This decomposition requires only the trial identifier for each patient, which is available to PRO-ACT data contributors and regulatory agencies but is not included in the public release. The analysis code required to perform this decomposition is publicly available at the repository listed in the Data Availability section.

A second direction is the application of the entropy-trajectory approach to other ordinal outcome scales (UPDRS, ADAS-Cog, EDSS, HAM-D, PANSS) using existing clinical trial databases. If the same pattern holds (treatment-arm entropy divergence invisible to total-score slopes), the finding generalizes beyond ALS to a systematic analytic failure mode across neurology and psychiatry.

## 5 Conclusion

Computing Shannon entropy from item-level ALSFRS score distributions, stratified by treatment arm, reveals systematic divergence between drug-treated and placebo populations across all four functional domains in the PRO-ACT database. In the Fine Motor domain, the active arm’s entropy peak is delayed by five months relative to placebo. This divergence is model-independent, visible in the raw entropy trajectories, and confirmed by permutation testing at *p <* 0.001 (7.5 standard deviations above the null mean). The quantity that differs between arms, the shape and timing of the population’s distributional evolution, is not representable in the total-score linear-slope framework used to evaluate these trials. Whether this signal reflects treatment effects, compositional artifacts from pooling heterogeneous trials, or both cannot be resolved from the anonymized public database. What can be determined is that the standard endpoint discards information that is empirically non-empty: the distributional structure it assumes to be uninformative contains statistically significant between-arm divergence.

## Data availability

Analysis code is available at https://github.com/amr28693/als_arm_entropy. Data were obtained from the PRO-ACT database (https://ncri1.partners.org/ProACT) under the standard data use agreement.

## Acknowledgments

Data used in the preparation of this article were obtained from the Pooled Resource Open-Access ALS Clinical Trials (PRO-ACT) Database. The data available in the PRO-ACT Database have been volunteered by PRO-ACT Consortium members.

## Supplementary Materials

**Figure S1:**
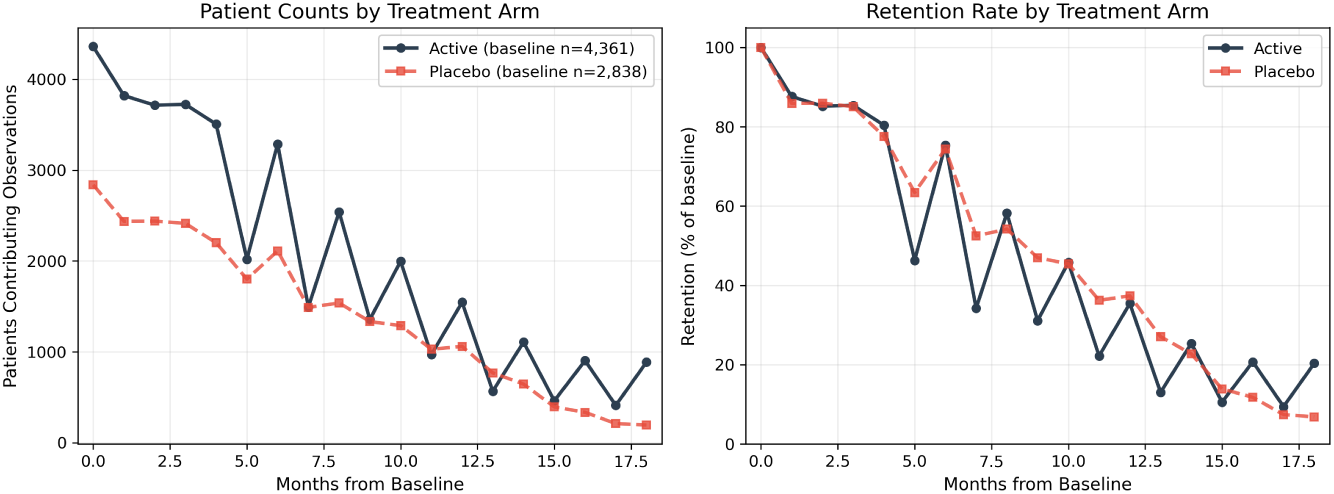
Attrition profile. Patient attrition by treatment arm. Left: number of patients contributing observations at each monthly time point. Right: retention as a percentage of baseline enrollment. The non-monotonic (sawtooth) pattern reflects heterogeneous visit schedules across the constituent trials pooled in the PROACT database; it does not indicate patients dropping out and re-entering. Both arms show progressive overall attrition over the 18-month observation window, with similar retention rates through month 5 and increasing divergence thereafter. The placebo arm retains a higher percentage of its baseline enrollment than the active arm from month 5 onward; this differential retention is addressed in the Limitations.

**Table S1:** Patient attrition by treatment arm. Number of patients contributing observations at each monthly time point by treatment arm. Month-0 counts (4,361 Active; 2,838 Placebo) are lower than total unique patients with arm labels (4,581 Active; 2,931 Placebo) because some patients’ earliest observations round to month 1 or later. The non-monotonic pattern reflects heterogeneous visit schedules across the pooled constituent trials.

| Month | $n_{\text{Active}}$ | $n_{\text{Placebo}}$ | % Active | % Placebo |
| --- | --- | --- | --- | --- |
| 0 | 4,361 | 2,838 | 100.0 | 100.0 |
| 1 | 3,821 | 2,437 | 87.6 | 85.9 |
| 2 | 3,715 | 2,439 | 85.2 | 85.9 |
| 3 | 3,724 | 2,414 | 85.4 | 85.1 |
| 4 | 3,506 | 2,200 | 80.4 | 77.5 |
| 5 | 2,020 | 1,800 | 46.3 | 63.4 |
| 6 | 3,286 | 2,111 | 75.3 | 74.4 |
| 7 | 1,492 | 1,489 | 34.2 | 52.5 |
| 8 | 2,539 | 1,538 | 58.2 | 54.2 |
| 9 | 1,357 | 1,333 | 31.1 | 47.0 |
| 10 | 1,996 | 1,289 | 45.8 | 45.4 |
| 11 | 969 | 1,028 | 22.2 | 36.2 |
| 12 | 1,545 | 1,060 | 35.4 | 37.4 |
| 13 | 568 | 768 | 13.0 | 27.1 |
| 14 | 1,107 | 645 | 25.4 | 22.7 |
| 15 | 461 | 394 | 10.6 | 13.9 |
| 16 | 903 | 334 | 20.7 | 11.8 |
| 17 | 411 | 210 | 9.4 | 7.4 |
| 18 | 887 | 194 | 20.3 | 6.8 |

**Table S2:** Sensitivity to minimum-*n* threshold. Sensitivity of divergence metrics to the minimum observation threshold per item per month. All three thresholds yield the same 19 common time points (months 0–18). The five-month Fine Motor peak delay and all domain-level significance patterns are unchanged. The min-*n* = 20 results are numerically identical to min-*n* = 30, indicating that no item-month-arm bins in the dataset contain between 20 and 29 observations.

| Metric | min-$n = 20$ | min-$n = 30$ | min-$n = 50$ |
| --- | --- | --- | --- |
| Total IAD | 4.476 | 4.476 | 4.593 |
| Total $p$-value | <0.001 | <0.001 | <0.001 |
| Total $\sigma$ from null | 7.5 | 7.5 | 7.8 |
| Fine Motor IAD | 0.545 | 0.545 | 0.508 |
| Fine Motor $p$-value | 0.001 | 0.001 | 0.001 |
| Fine Motor peak (Active) | 13 | 13 | 13 |
| Fine Motor peak (Placebo) | 8 | 8 | 8 |
| Respiratory IAD | 3.062 | 3.062 | 3.168 |
| Respiratory $p$-value | <0.001 | <0.001 | <0.001 |
| Bulbar IAD | 0.588 | 0.588 | 0.641 |
| Bulbar $p$-value | 0.263 | 0.263 | 0.195 |
| Gross Motor IAD | 0.282 | 0.282 | 0.277 |
| Gross Motor $p$-value | 0.579 | 0.579 | 0.600 |

